# Autoimmune conditions following mRNA (BNT162b2) and inactivated (CoronaVac) COVID-19 vaccination: a descriptive cohort study among 1.1 million vaccinated people in Hong Kong

**DOI:** 10.1101/2021.10.21.21265314

**Authors:** Xue Li, Le Gao, Xinning Tong, Vivien K.Y. Chan, Celine S.L. Chui, Francisco T.T. Lai, Carlos K.H. Wong, Eric Y.F. Wan, Esther W.Y. Chan, Kui Kai Lau, Chak Sing Lau, Ian C.K. Wong

## Abstract

**Background:** Concerns regarding the autoimmune safety of COVID-19 vaccines may negatively impact vaccine uptake. We aimed to describe the incidence of autoimmune conditions following BNT162b2 and CoronaVac vaccination and compare these with age-standardized incidence rates in non-vaccinated individuals.

**Methods:** This is a descriptive cohort study conducted in public healthcare service settings. Territory-wide longitudinal electronic medical records of Hong Kong Hospital Authority users (≥16 years) were linked with COVID-19 vaccination records between February 23, 2021 and June 30, 2021. We classified participants into first/second dose BNT162b2 groups, first/second dose CoronaVac groups and non-vaccinated individuals for incidence comparison. The study outcomes include hospitalized autoimmune diseases (16 types of immune-mediated diseases across six body systems) within 28 days after first and second dose of vaccination. Age-standardized incidence rate ratios (IRRs) with exact 95% confidence intervals (CIs) were estimated using Poisson distribution.

**Results:** This study included around 3.9 million Hong Kong residents, of which 1,122,793 received at least one dose of vaccine (BNT162b2: 579,998; CoronaVac: 542,795), and 721,588 completed two doses (BNT162b2: 388,881; CoronaVac: 332,707). Within 28 days following vaccination, cumulative incidences for all autoimmune conditions were below 9 per 100,000 persons, for both vaccines and both doses. None of the age-standardized incidence rates were significantly higher than the non-vaccinated individuals, except for an observed increased incidence of hypersomnia following the first dose of BNT162b2 (standardized IRR: 1.47; 95% CI: 1.10–1.94).

**Conclusions:** Autoimmune conditions requiring hospital care are rare following mRNA and inactivated virus COVID-19 vaccination with similar incidence to non-vaccinated individuals. The association between first dose BNT162b2 vaccination and immune-related sleeping disorders requires further research. Population-based robust safety surveillance is essential to detect rare and unexpected vaccine safety events.

**Funding:** Research Grant from the Food and Health Bureau, the Government of the Hong Kong Special Administrative Region (Ref. No. COVID19F01).

## Introduction

There has been a long-standing debate about vaccination and its potential to trigger autoimmune diseases (AID). For example, the influenza vaccine and Guillain-Barré syndrome^1^, the Measles-Mumps-Rubella vaccine and thrombocytopenic purpura^2^. Although causality is not fully established, the fear of new onset or recurrence of AID may contribute to vaccine hesitancy and negatively impact vaccine uptake^3^. Currently, several vaccines using different technology platforms (mRNA, inactivated virus, protein subunit and viral vector) have been developed and authorized for emergency use against COVID-19 worldwide^4^. The characteristics of these vaccine platforms could result in different levels of neutralizing antibodies, T-cell response, and avoidance/occurrence of immune-mediated diseases.

Autoimmune safety events reported in Phase II/III trials, such as Bell’s Palsy in mRNA vaccine (Moderna)^5^ and transverse myelitis in viral vector vaccine (AstraZeneca (ChAdOx1))^6^, were uncommon and have been labeled in the prescribing information. However, since the launch of global vaccination against COVID-19, several case reports and analyses have described autoimmune conditions following vaccination. In a prospective cohort analysis, Simpson et al. implied an association between viral vector vaccine (AstraZeneca (ChAdOx1)) and the development of immune thrombocytopenia^7^. Wan et al. suggested the safety signal of Bell’s palsy with inactivated vaccine (CoronaVac) in a nested case-control and case series study^8^. Case reports or case series of Guillain-Barré syndrome^9-12^, cutaneous vasculitis^13^, reactive arthritis^14^ and immune-medicated disease flare^15^ from both mRNA and inactivated virus platforms are continuously emerging, and the list is not exclusive. Given the ongoing COVID-19 pandemic and the likelihood of routine vaccination for infection control, it is of timely importance to assess the autoimmune safety of COVID-19 vaccines in real-world settings.

Hong Kong (HK) is among the few jurisdictions in the world that has approved the emergency use of COVID-19 vaccines from two different technology platforms: inactivated virus vaccine, CoronaVac, from Sinovac Biotech (HK) Limited and mRNA vaccine, BNT162b2, from Fosun-BioNTech (equivalent to Pfizer-BioNTech). The rollout of the public-funded mass vaccination program began with CoronaVac on February 23, 2021 and was closely followed with BNT162b2 on March 12, 2021. By June 30, 2021, more than 3.73 million doses of COVID-19 vaccine had been administered alongside the implementation of population-based surveillance program for safety monitoring. In this study, we analyzed the territory-wide electronic medical records (EMRs) database and summarized the incidence of AID across the predefined disease spectrum among CoronaVac and BNT162b2 recipients in HK. We assessed the population-level risk of AID following mRNA and inactivated virus COVID-19 vaccination to inform vaccination decisions.

## Methods

### Data sources

We obtained population-based EMRs from the Hospital Authority (HA) and vaccination records from the Department of Health (DH) of HK Government to conduct this study. The HA manages all public hospitals and clinics in HK and provides publicly funded health services to all eligible HK residents (>7 million). The EMRs database managed by HA is centralized from the territory-wide clinical management system of 42 public hospitals with high population coverage, representativeness, and coding accuracy^16-19^. DH, the government health adviser and agency that executes health-related policies and statutory functions, manages the COVID-19 vaccination records of all HK residents. EMRs from the HA were linked with the DH vaccination records using de-identified non-reversible pseudo-ID to protect patient privacy.

### Study design and population

This was a population-based descriptive cohort study. The study population consisted of all Hong Kong residents ≥16 years who had ever used the HA service (emergency admission, general or specialist outpatient clinic visit, or hospital admission) between January 1, 2018 and June 30, 2021. DH vaccination records between February 23 and June 30, 2021 were linked to the HA database to confirm vaccination status. Following the vaccination record linkage, we generated pseudo index dates for the non-vaccinated individuals to facilitate cohort follow-up. Within each age and sex group, we matched each vaccine recipient with non-vaccinated individuals at maximum ratio (the number of vaccine recipients divided by the number of non-vaccinated individuals in each age-sex strata) and assigned the vaccination date as the pseudo index date for the non-vaccinated individuals. We treated first dose and second dose as independent episodes and conducted the non-vaccinated group matching separately.

We followed up these individuals from the index date (date of first dose vaccination, date of second dose vaccination, or age-sex matched pseudo index date for non-vaccinated individuals) to the date of event of interest, date of registered death, 28 days after index date, or the end date of data availability (June 30, 2021), whichever was earlier. To differentiate whether the event was completely related to the first dose, in the first dose analysis, we also censored the follow-up one day before the second dose, if any. The record linkage, matching procedure and cohort follow-up is illustrated in Supplementary Figure 1.

### Outcome measures

We assessed the incidence of hospital admission related to a spectrum of 16 pre-specified AID, grouped by body systems including: 1) immune-mediated cardiovascular diseases (Kawasaki disease, single organ cutaneous vasculitis); 2) organ specific immune-mediated endocrine disorder (subacute thyroiditis); 3) immune-mediated hematological diseases (anti-phospholipid antibody syndrome, idiopathic thrombocytopenia); 4) immune-mediated multisystem diseases (Sjogren syndrome, systemic lupus erythematosus); 5) immune-mediated musculoskeletal diseases (acute aseptic arthritis, reactive arthritis, rheumatoid arthritis, psoriatic arthritis, spondyloarthritis); and 6) immune-mediated neurological disorders (acute disseminated encephalomyelitis, Guillain-Barré syndrome, narcolepsy and related disorders, and transverse myelitis). We used the International Classification of Diseases, Ninth Revision, Clinical Modification (ICD-9-CM) diagnosis codes for outcomes identification (Supplementary Table 1). We defined each individual outcome as an incident case if this was the patient’s first relevant diagnosis (primary diagnosis) in the inpatient setting during the 28 days of follow-up. Patients with a diagnosis history within 365 days before the index date were excluded from the study cohort. The same procedure was conducted for first dose, second dose and non-vaccinated groups.

### Main analysis

We reported two age-standardized health situation measurements as the main outcome of this study. First, we described the 28-day cumulative incidence (per 100,000 persons) of each condition, using 5-year age band weightings (16–19, 20– 24, 25–29, 30–34, 35–39, 40–44, 45–49, 50–54, 55–59,60–64, 65–69, 70–74, 75– 79, 80–84 and ≥85 years) based on the HK 2021 mid-year population census. Second, to account for the follow-up time differences among groups, we reported the age-standardized incidence rate (per 100,000 person-years) using the indirect standardization method. We treated the non-vaccinated group as an internal reference group and calculated the proportion of follow-up time (in person-years) in each age strata (16–19, 20–24, 25–29, 30–34, 35–39, 40–44, 45–49, 50–54, 55–59, 60–64, 65–69, 70–74, 75–79, 80–84 and ≥85 years) for incidence standardization. For each condition, we used observed number of cases, the follow-up time and proportion of person-years in each age strata to calculate age-specific crude incidence rate and aggregated the age-standardized incidence rate. We further estimated the expected cases in each group and calculated the standardized incidence rate ratios (IRRs) with exact 95% confidence intervals (CIs) using Poisson distribution.^20,21^

### Additional analyses

We conducted several additional analyses. First, in order to exclude the possibility that the autoimmune conditions were related to SARS-CoV-2 infection, we removed individuals who had positive SARS-CoV-2 PCR tests prior to June 30, 2021 and replicated the main analysis. Second, considering that some of the autoimmune manifestations, e.g., Guillain-Barré syndrome, may take longer than 28 days to develop, we removed the 28-day follow-up requirement in the second dose analysis and extended the observation period to the end date of data availability. Third, for the estimation of standardized IRRs, we further extended the case definition using both primary and secondary diagnoses at discharge, in consideration of potential different coding practices by clinicians. In addition, we conducted an ecological study to complement the main analysis. We assumed that the COVID-19 vaccination program (implemented from March to June 2021) will not impact the natural occurrence of AID at population-level and the incidence would be stable over the years within the study cohort. Hence, we illustrated a four-month (March-June) cumulative incidence trend for each AID between 2018 and 2021. The observed number of incident cases were divided by the total number of HA active patients (≥16 years) in the corresponding year to calculate the cumulative incidence. Age-standardization was also conducted based on the HK 2021 mid-year population census. All analyses were conducted under Microsoft^®^ Excel^®^ and R 4.0.3 (with R code available at https://github.com/legao513/Auto_immune_descriptive/blob/main/analysis_script_0927.R).

### Ethics

Ethical approval for this study was granted by the Institutional Review Board of the University of Hong Kong/ Hospital Authority Hong Kong West Cluster (UW 21-149 and UW 21-138); and the Department of Health Ethics Committee (LM 21/2021). Patient identification was anonymized from HA and DH databases and patient consent was not required.

## Results

We obtained the EMRs of 3,946,550 HA active patients with affirmed vaccination status. After age-sex matching with the non-vaccinated group, 1,122,793 vaccine recipients (BNT162b2: 579,998; CoronaVac: 542,795) were included in the first dose analysis, and 721,588 vaccine recipients (BNT162b2: 388,881; CoronaVac: 332,707) were included in the second dose analysis with follow-up until censoring (Supplementary Figure 1). 63.0% in the first dose analysis and 71.9% in the second dose analysis had the injection on or before June 3, 2021 with the full 28 days observation from vaccination date to the study end date (June 30, 2021). Compared with non-vaccinated individuals, vaccine recipients were younger, more likely to be males, and less likely to have pre-existing chronic diseases. Detailed baseline characteristics of the study cohorts are summarized in Table 1.

**Table 1.** Baseline characteristics and pre-existing comorbidities of study cohorts.

|  | First dose |  |  | Second dose |  |  |
| --- | --- | --- | --- | --- | --- | --- |
|  | None | BNT162b2 | CoronaVac | None | BNT162b2 | CoronaVac |
| N | 2816133 | 579998 | 542795 | 1892783 | 388881 | 332707 |
| Male (%) | 1183236 (42.0) | 275965 (47.6) | 266681 (49.1) | 831868 (43.9) | 190562 (49.0) | 175487 (52.7) |
| Age (mean (SD)) | 55.67 (19.17) | 46.72 (15.06) | 54.01 (13.85) | 56.81 (19.27) | 47.49 (14.88) | 54.67 (13.97) |
| Charlson Comorbidity Index (mean (SD)) | 0.35 (0.91) | 0.10 (0.41) | 0.15 (0.49) | 0.37 (0.93) | 0.10 (0.41) | 0.15 (0.49) |
| Myocardial infarction | 16935 (0.6) | 617 (0.1) | 891 (0.2) | 12229 (0.6) | 418 (0.1) | 548 (0.2) |
| Congestive Heart Failure | 32695 (1.2) | 425 (0.1) | 912 (0.2) | 24414 (1.3) | 302 (0.1) | 581 (0.2) |
| Peripheral vascular disease | 7525 (0.3) | 249 (0.0) | 362 (0.1) | 5631 (0.3) | 175 (0.0) | 240 (0.1) |
| Cerebrovascular disease | 111165 (3.9) | 4309 (0.7) | 7350 (1.4) | 80569 (4.3) | 2964 (0.8) | 4611 (1.4) |
| Stroke or systemic embolism | 37828 (1.3) | 1226 (0.2) | 2197 (0.4) | 27494 (1.5) | 852 (0.2) | 1360 (0.4) |
| Asthma | 27317 (1.0) | 3665 (0.6) | 3309 (0.6) | 18798 (1.0) | 2468 (0.6) | 2043 (0.6) |
| Chronic obstructive pulmonary disease | 58132 (2.1) | 4778 (0.8) | 5343 (1.0) | 41664 (2.2) | 3244 (0.8) | 3365 (1.0) |
| Dementia | 11122 (0.4) | 103 (0.0) | 149 (0.0) | 8477 (0.4) | 69 (0.0) | 105 (0.0) |
| Diabetes without chronic complication | 351241 (12.5) | 27787 (4.8) | 41633 (7.7) | 245849 (13.0) | 18561 (4.8) | 25251 (7.6) |
| Diabetes with chronic complication | 20940 (0.7) | 871 (0.2) | 1302 (0.2) | 14766 (0.8) | 594 (0.2) | 795 (0.2) |
| Chronic renal failure | 38702 (1.4) | 1155 (0.2) | 2048 (0.4) | 28288 (1.5) | 778 (0.2) | 1297 (0.4) |
| Mild liver disease | 4056 (0.1) | 231 (0.0) | 324 (0.1) | 2846 (0.2) | 158 (0.0) | 188 (0.1) |
| Moderate-severe liver disease | 2871 (0.1) | 141 (0.0) | 188 (0.0) | 1980 (0.1) | 95 (0.0) | 108 (0.0) |
| Rheumatoid arthritis and other inflammatory polyarthropathies | 10584 (0.4) | 813 (0.1) | 873 (0.2) | 7010 (0.4) | 479 (0.1) | 456 (0.1) |
| Malignancy | 75657 (2.7) | 4682 (0.8) | 5220 (1.0) | 52132 (2.8) | 3054 (0.8) | 3107 (0.9) |
| Metastatic solid tumor | 14757 (0.5) | 348 (0.1) | 418 (0.1) | 10256 (0.5) | 226 (0.1) | 234 (0.1) |

Note: P-values for vaccinated and non-vaccinated groups comparison all <0.001.

### Main analysis

Within 28 days after the index date, autoimmune conditions requiring hospitalization were generally rare (cumulative incidence all below 9 per 100,000 persons) among vaccine recipients and non-vaccinated individuals (Figure 1). After the first dose, most of the analyzed immune-mediated cardiovascular, endocrine, hematological, nervous, and multisystem diseases had a cumulative incidence lower than 1 per 100,000 persons for both BNT162b2 and CoronaVac recipients, translating to fewer than 10 cases per 1 million of first dose administration (Table 2). Several autoimmune conditions following immunization are extremely rare and zero cases were recorded in our database for acute disseminated encephalomyelitis, Kawasaki disease, subacute thyroiditis, and transverse myelitis for BNT162b2 and CoronaVac and also Guillain-Barré syndrome, single organ cutaneous vasculitis for CoronaVac. The incidence of reactive arthritis and narcolepsy-related disorders were relatively higher compared to other AIDs, with cumulative incidence ranging between 4.12 and 8.35 per 100,000 persons among first dose vaccine recipients (Table 2). Similarly, the majority of the autoimmune conditions following the second dose also had a 28-day cumulative incidence of less than 1 per 100,000 persons (Figure 1). Reactive arthritis, narcolepsy-related disorders and thrombocytopenia appeared to have a numerically higher incidence than the corresponding incidence reported after the first dose.

**Figure 1.**
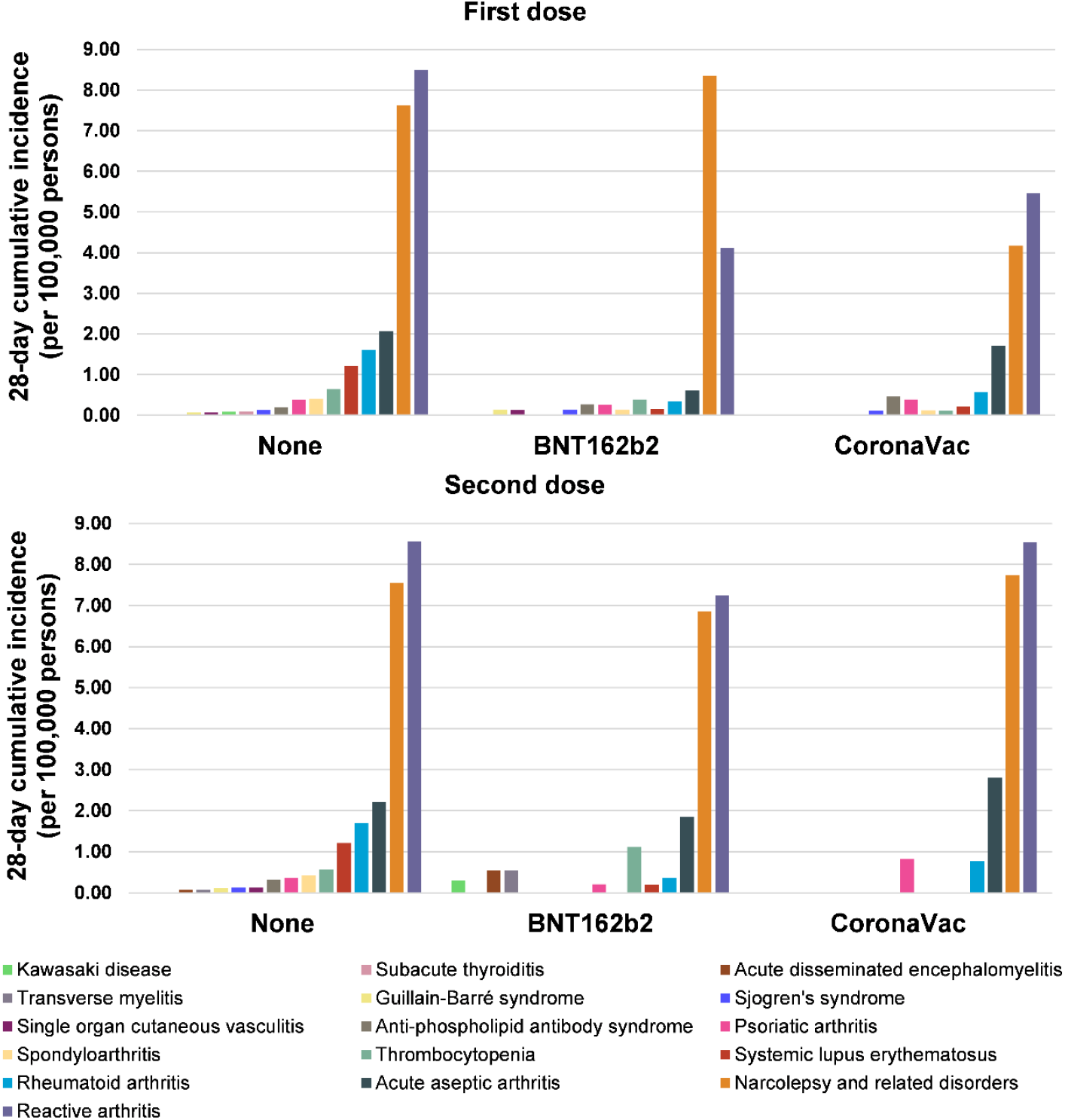
Cumulative incidence of hospitalized autoimmune diseases among vaccine recipients and non-vaccinated individuals.

**Table 2.** 28-day cumulative incidence (per 100,000 persons) of autoimmune conditions among vaccine recipients and non-vaccinated individuals.

| Body system | Disease | First dose |  |  | Second dose |  |  |
| --- | --- | --- | --- | --- | --- | --- | --- |
|  |  | None | BNT162b2 | CoronaVac | None | BNT162b2 | CoronaVac |
| Cardiovascular system | Kawasaki disease | 0.080 | 0.000 | 0.000 | 0.00 | 0.29 | 0.00 |
|  | Single organ cutaneous vasculitis | 0.066 | 0.13 | 0.00 | 0.12 | 0.00 | 0.00 |
| Endocrine system | Subacute thyroiditis | 0.090 | 0.00 | 0.00 | 0.00 | 0.00 | 0.00 |
| Hematological system | Anti-phospholipid antibody syndrome | 0.19 | 0.27 | 0.46 | 0.32 | 0.00 | 0.00 |
|  | Thrombocytopenia | 0.64 | 0.39 | 0.11 | 0.57 | 1.11 | 0.00 |
| Multisystem | Sjogren's syndrome | 0.13 | 0.14 | 0.11 | 0.12 | 0.00 | 0.00 |
|  | Systemic lupus erythematosus | 1.21 | 0.15 | 0.22 | 1.21 | 0.20 | 0.00 |
| Musculoskeletal system | Acute aseptic arthritis | 2.07 | 0.61 | 1.71 | 2.21 | 1.85 | 2.80 |
|  | Reactive arthritis | 8.49 | 4.12 | 5.46 | 8.56 | 7.24 | 8.53 |
|  | Rheumatoid arthritis | 1.60 | 0.34 | 0.57 | 1.69 | 0.36 | 0.77 |
|  | Psoriatic arthritis | 0.38 | 0.26 | 0.39 | 0.37 | 0.20 | 0.82 |
|  | Spondyloarthritis | 0.40 | 0.14 | 0.12 | 0.42 | 0.00 | 0.00 |
| Nervous system | Acute disseminated encephalomyelitis | 0.00 | 0.00 | 0.00 | 0.071 | 0.55 | 0.00 |
|  | Guillain-Barré syndrome | 0.062 | 0.14 | 0.00 | 0.10 | 0.00 | 0.00 |
|  | Narcolepsy and related disorders | 7.62 | 8.35 | 4.17 | 7.55 | 6.86 | 7.74 |
|  | Transverse myelitis | 0.00 | 0.00 | 0.00 | 0.07 | 0.55 | 0.00 |
| All-cause death | Death | 60.47 | 3.29 | 4.54 | 63.18 | 6.83 | 3.98 |

When considering the follow-up time and measuring the disease occurrence using standardized incidence rate, we did not detect a significantly increased incidence rate of all the interested autoimmune conditions among the vaccine recipients (Figure 2 first dose analysis; Figure 3 second dose analysis), except narcolepsy and related disorders among first dose BNT162b2 recipients (standardized incidence: 220.56 versus 136.29 per 100,000 person-years; standardized IRR: 1.47; 95% CI: 1.10–1.94). The age-standardized all-cause mortality was considerably lower in the vaccine groups, suggesting that individuals who received the vaccine might be healthier than the non-vaccinated. Median-time to event since first or second dose of vaccination ranged between 3 and 28 days (Supplementary Table 2).

**Figure 2.**
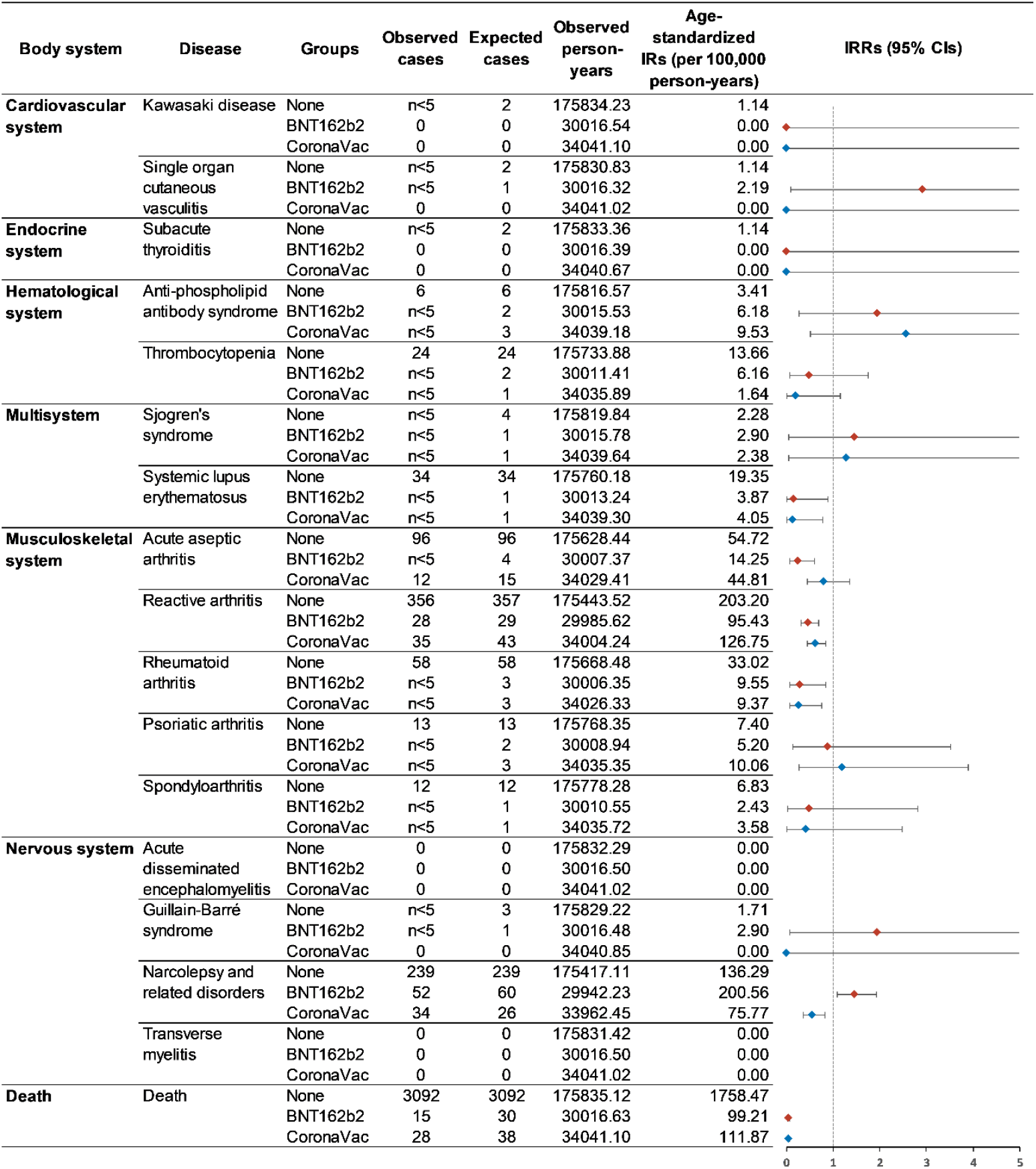
Standardized incidence rates of autoimmune conditions among first dose vaccine recipients and non-vaccinated individuals. Note: The line for CIs without a cap means the upper CI is greater than 5. Abbreviations: CIs, confidence intervals, IRs, incidence rates, IRRs, incidence rate ratios.

**Figure 3.**
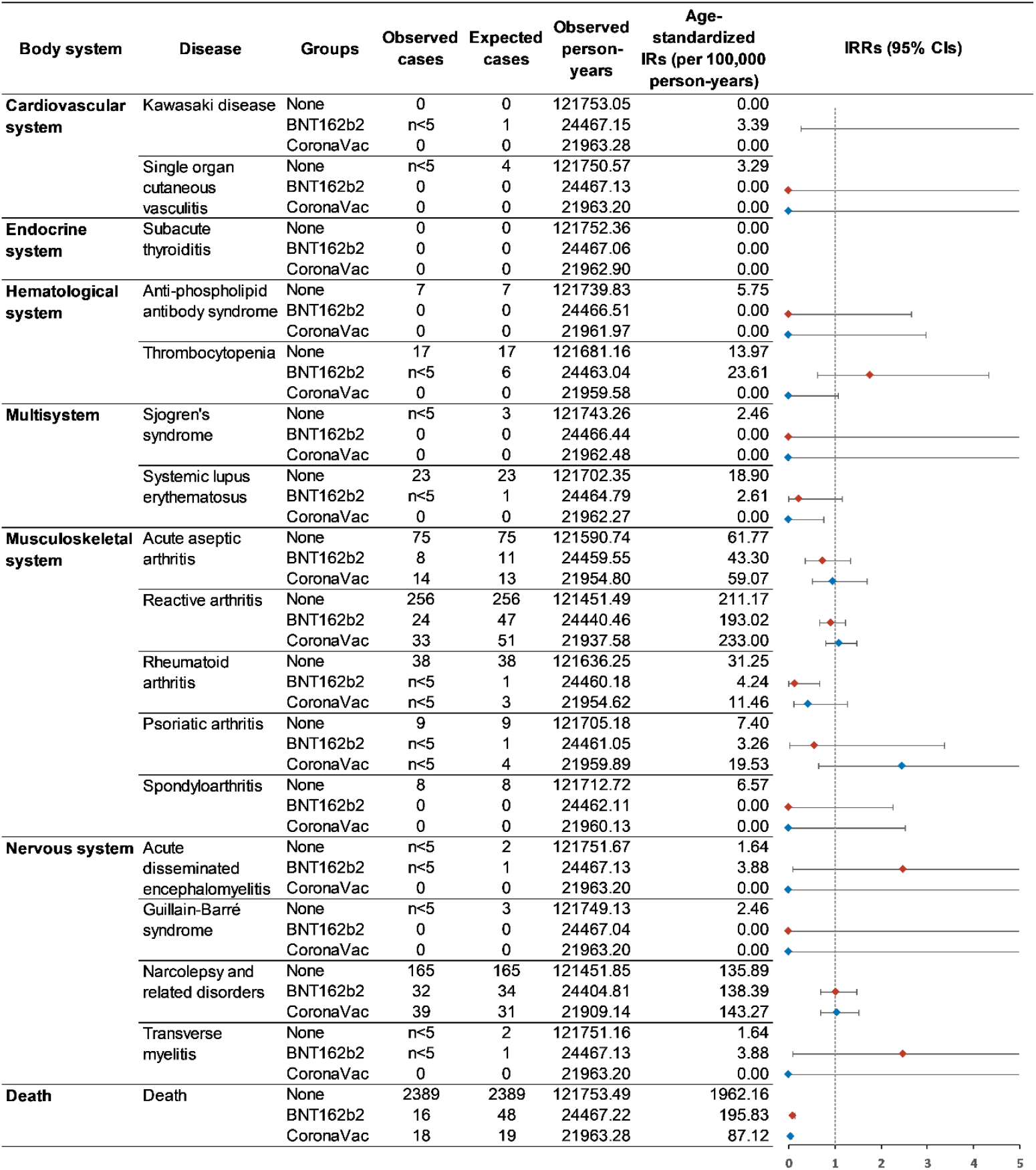
Standardized incidence rates of autoimmune conditions among second dose vaccine recipients and non-vaccinated individuals. Note: The line for CIs without a cap means the upper CI is greater than 5. Abbreviations: CIs, confidence intervals, IRs, incidence rates, IRRs, incidence rate ratios.

### Additional analyses

In total, we removed 10,185 individuals who had previous SARS-CoV-2 infection before our study end date, and the results were consistent with the main analysis (Supplementary Figure 2 and Supplementary Figure 3). After removing the 28-day follow-up time restriction in the second dose analysis, we observed that there were more autoimmune conditions with non-significant IRRs greater than one, including Sjogren’s syndrome, acute disseminated encephalomyelitis and transverse myelitis in both BNT162b2 and CoronaVac groups, psoriatic arthritis in CoronaVac group, and narcolepsy and related disorders in the BNT162b2 group (Supplementary Figure 4). When using both primary and secondary diagnoses for case definition, the standardized IRRs for all disease groups yielded consistent results as the main analysis (Supplementary Figure 5 and Supplementary Figure 6). The tendency for increased risk of narcolepsy and related disorder among first dose BNT162b2 recipients remained but became non-significant (standardized incidence: 242.26 versus 197.88 per 100,000 person-years standardized IRR: 1.23; 95% CI: 0.95– 1.58). In the ecological analysis of 3.9 million HA active patients, we observed a relatively stable trend of a four-month cumulative incidence between 2018 and 2021 for all autoimmune conditions analyzed (Supplementary Figure 7).

## Discussion

Hong Kong is among the ten jurisdictions in the world implementing both mRNA and inactivated virus vaccines for emergency use, which allows us to analyze the autoimmune safety performance of the two technology platforms in one population cohort. In this territory-wide descriptive cohort study following more than 1 million people with BNT162b2 and CoronaVac vaccination in HK, we found that autoimmune conditions that need inpatient management are rare, with a comparable incidence rate with non-vaccinated individuals. Under the circumstance of emerging case reports on autoimmune conditions following COVID-19 vaccination, this study provides population-level perspectives on autoimmune safety, with the potential to enhance vaccine confidence among the public.

Among the autoimmune conditions analyzed in this study, published case reports focused mainly on Guillain-Barré syndrome, immune thrombocytopenia, and cutaneous vasculitis^7,9-13,22^. Because of the low incidence, the observed number of these three conditions are all less than 5 among our 1.2 million vaccine recipient records. Within 28 days after vaccination, most of the standardized incidence rates were not significantly higher than in non-vaccinated individuals, and we were unable to detect major safety signals for these conditions for both vaccines. The wide range of median time to event following vaccination indicates the heterogeneity of immune response in different body systems. In the additional analysis that removed the requirement of 28 days follow-up, we observed more diseases with an IRR greater than one although they did not reach significant levels. This might suggest that some autoimmune manifestations may take longer than 28 days to develop and the risk window for vaccine safety monitoring needs thoughtful adjustment. The non-significant results shown in this study reflect the natural rarity of AID, which proposes a huge challenge for signal detection and association evaluation. The World Health Organization has called for pharmacovigilance preparedness for COVID-19 vaccines in all countries.^23^ Our study highlights that, for rare safety events monitoring, it is particularly imperative to establish a population-based, real-time surveillance system through multiple stakeholder collaboration and capacity building.

Similar to the observations from other COVID-19 vaccine safety studies, individuals who received the vaccines are generally healthier with fewer comorbidities than the general population^24,25^. In HK, the rollout of the COVID-19 vaccination program prioritized healthcare workers, personnel maintaining critical public services and care home residents at the initial stage^26^. Four months after the program commencement, 44% of the eligible HK population had already received the first vaccine dose while the uptake in care home residents was only 5%. This possibly explains the significantly lower incidence of systemic lupus erythematosus, reactive arthritis, and rheumatoid arthritis, and most pronouncedly, all-cause mortality, among vaccine recipients found in this study. Notably, this observation should not be misinterpreted as the vaccine lowering the risk of these diseases or preventing death. Due to the limited case numbers of each disease in this descriptive cohort study, we considered only age and follow-up time, not other confounding factors, when assessing autoimmune safety following COVID-19 vaccination. It is anticipated that more individuals with well-controlled chronic conditions, including the elderly, will be vaccinated given the government’s commitment of achieving optimal vaccine uptake and herd immunity. The number of rare safety outcomes could possibly accumulate, and further analytical epidemiological studies are highly encouraged to evaluate the association with comprehensive covariate adjustment.

With great interest, we observed a potential safety signal of narcolepsy and related disorders following the first dose of BNT162b2. The initial motivation for including narcolepsy in the list of interested AIDs was due to the widely reported increased risk of narcolepsy among Pandemrix (AS03 adjuvanted pandemic A/H1N1 influenza vaccine) recipients in European countries and Canada during the 2009 H1N1 pandemic^27-32^, which could contribute to vaccine hesitancy against COVID-19^33,34^. Considering the potential delay and miscoded diagnosis of narcolepsy from inpatient settings^35^. we used a series of ICD-9-CM codes (307.4x, 347.xx and 780.5x, Supplementary Table 1) to investigate a broader range of relevant sleeping disorders according to ACCESS - a project funded by the EMA for the safety monitoring of COVID-19 vaccines^36^. When breaking down the ICD-9-CM codes, we found that hypersomnia (ICD-9-CM: 780.53) was predominately coded but no case was coded as narcolepsy (ICD-9-CM: 347). Therefore, the increased incidence should not be attributed to narcolepsy, but mainly associated with hypersomnia. To date, no narcolepsy-related disorder case report is related to COVID-19 vaccination. There is a recent proof-of-concept study using wearable devices found an increased sleeping duration up to four days post-BNT162b2 vaccination^37^. The sleeping signal detected in this study encourages further investigation on the immunogenicity, mechanism and association between mRNA vaccine and sleeping disorders.

To the best of our knowledge, this is the first population-based study describing a spectrum of AIDs subsequent to exposure to BNT162b2 and CoronaVac vaccination. With the availability of territory-wide longitudinal EMRs, this study systematically reported the incidence of autoimmune conditions among vaccine recipients and non-vaccinated controls in the same period and within the same health system. This would avoid reporting bias from enhanced surveillance when using the historical background incidence as the control. There are also limitations in this study. Due to limited data sources, our case definition was restricted to autoimmune conditions that were severe enough to warrant hospitalization and inpatient care. Mild, self-limiting autoimmune conditions not requiring inpatient treatment are beyond the scope of this study. Similarly, our EMRs database does not include health information from the private sector, and information on other vaccination which may cause such diseases. Finally, case identification largely depended on ICD-9-CM diagnosis codes, which could cast some uncertainty on coding practices and affirmative diagnosis. However, previous studies using the EMRs database from the HA showed high coding accuracy^16-19^ and our additional analyses using both primary and secondary diagnosis would minimize this risk.

## Conclusions

Within 28 days after BNT162b2 and CoronaVac vaccination, autoimmune conditions requiring hospitalization are rare and similar to disease occurrence among the non-vaccinated population. The association between first dose BNT162b2 vaccination and immune-related sleeping disorders requires further research. Population-based active safety surveillance is essential to detect rare and unexpected adverse events and serves as a useful tool for future hypothesis testing epidemiology studies.

## Data Availability

Due to the regulation from the data provider, we won't be able to share raw data and proceed with the data included in this study.

## Acknowledgments

This study was funded by a research grant from the Food and Health Bureau, The Government of the Hong Kong Special Administrative Region (Ref. No. COVID19F01). We thank members of the Expert Committee on Clinical Events Assessment Following COVID-19 Immunization for case assessment and colleagues from the Drug Office of the Department of Health and from the Hospital Authority for providing vaccination and clinical data. We also thank Ms. Lisa Lam for proofreading the manuscript.

---

**Supplementary Figure 1.**
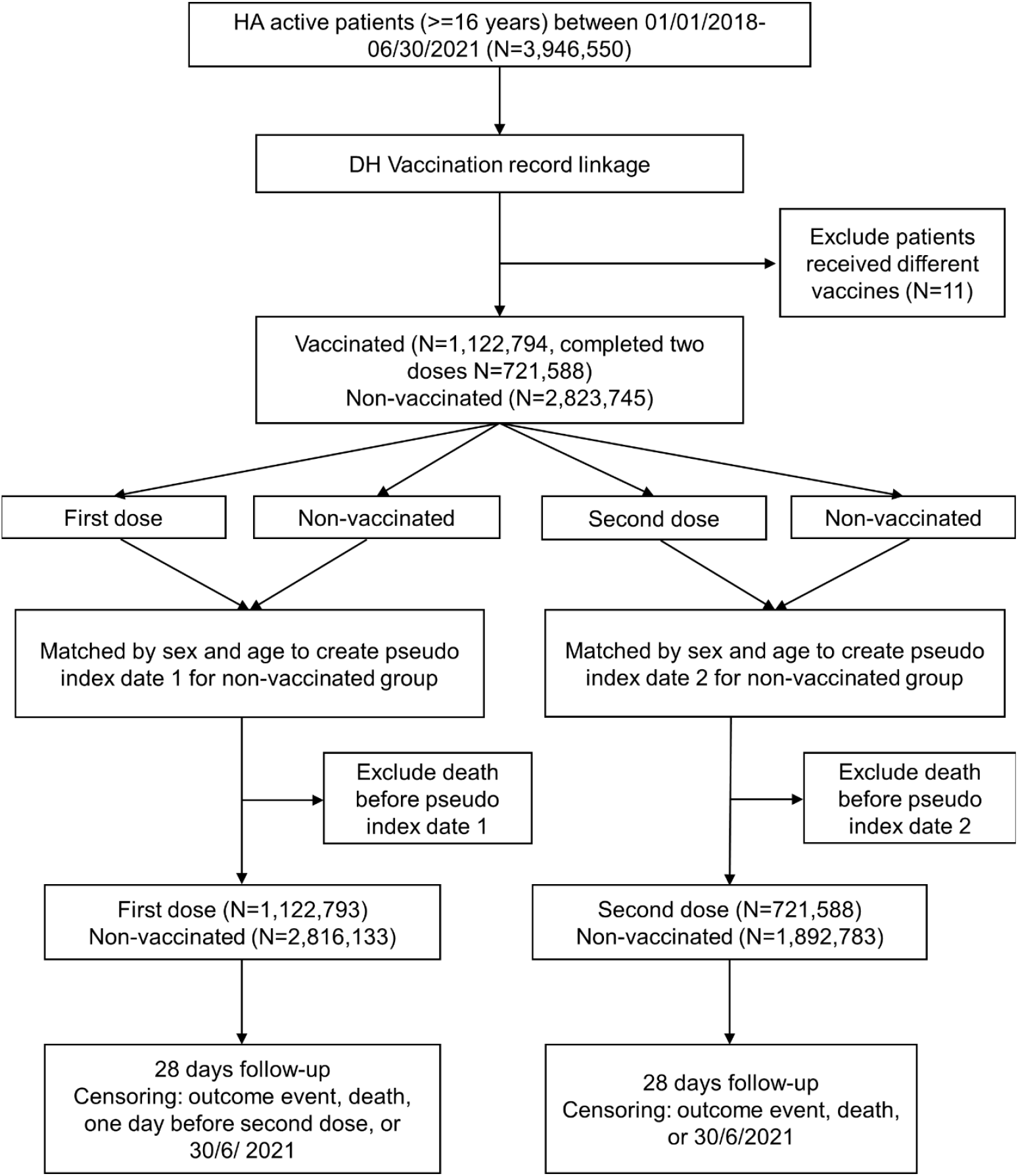
Study design and patient identification flow for main analysis. Abbreviations: HA, Hospital Authority; DH: Department of Health.

**Supplementary Figure 2.**
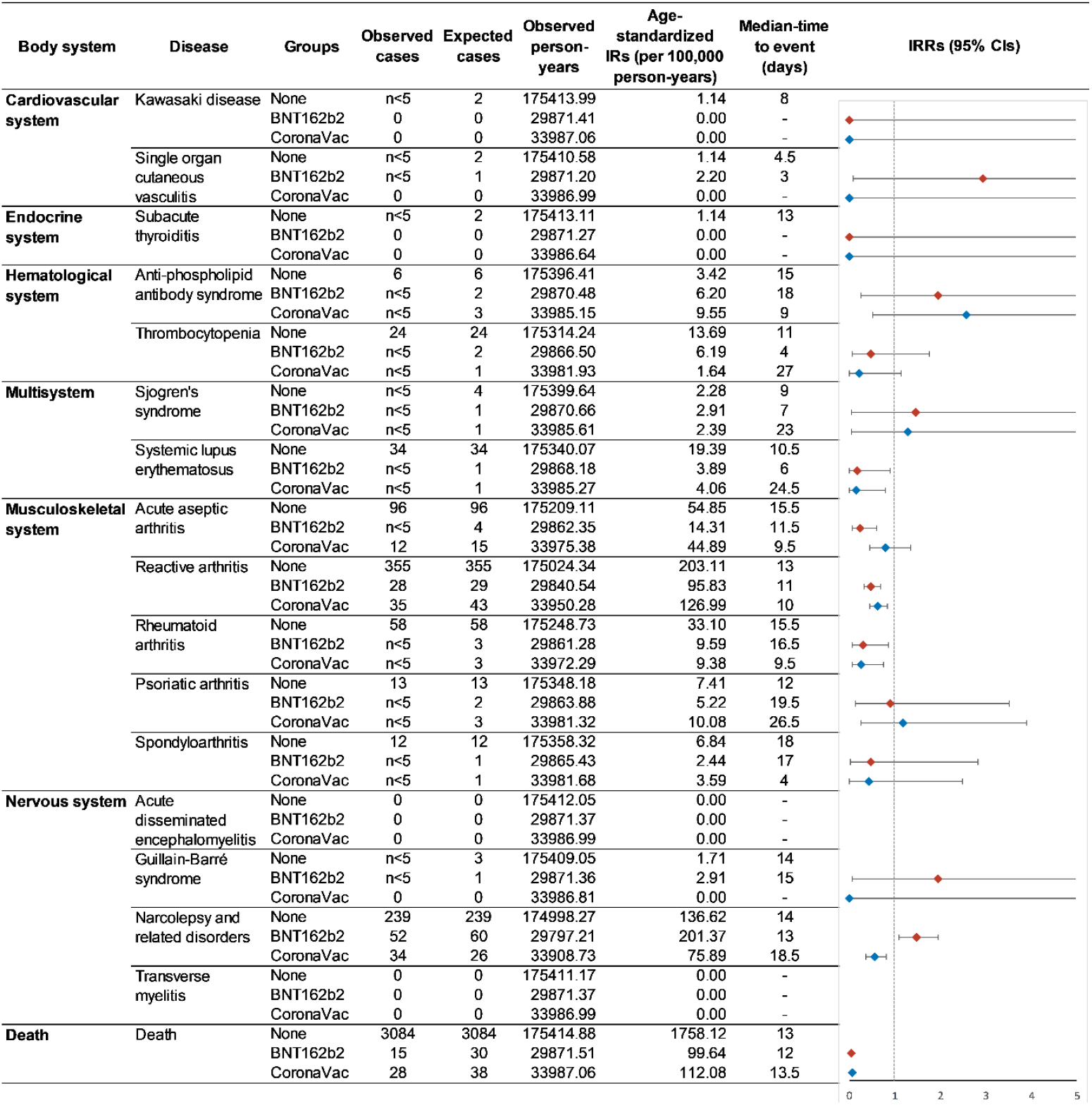
Standardized incidence rates of autoimmune conditions among first dose vaccine recipients and non-vaccinated individuals after removal of previous SARS-CoV-2 infection. Note: The line for CIs without a cap means the upper CI is greater than 5. Abbreviations: CIs, confidence intervals, IRs, incidence rates, IRRs, incidence rate ratios.

**Supplementary Figure 3.**
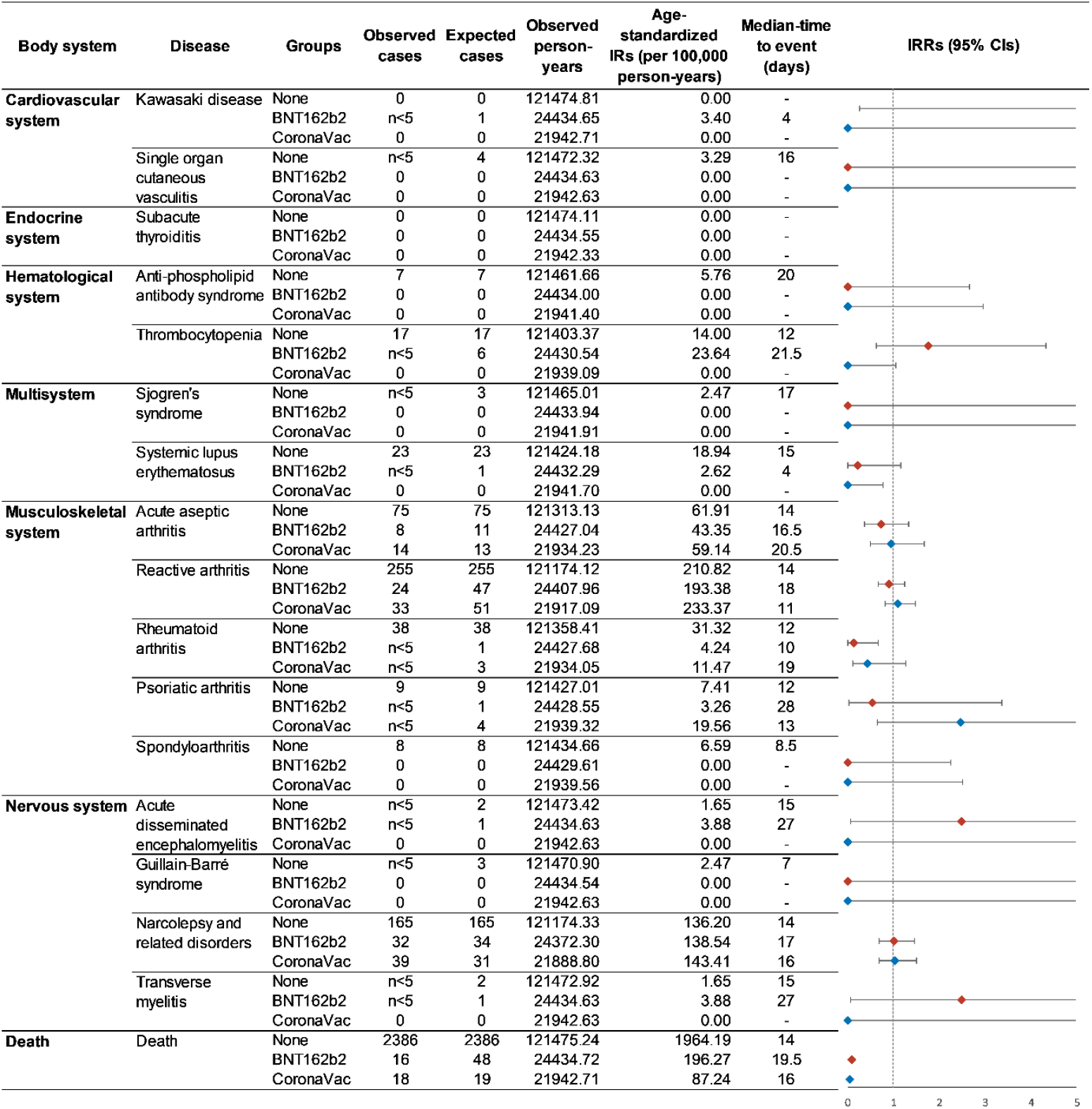
Standardized incidence rates of autoimmune conditions among second dose vaccine recipients and non-vaccinated individuals after removal of previous SARS-CoV-2 infection. Note: The line for CIs without a cap means the upper CI is greater than 5. Abbreviations: CIs, confidence intervals, IRs, incidence rates, IRRs, incidence rate ratios.

**Supplementary Figure 4.**
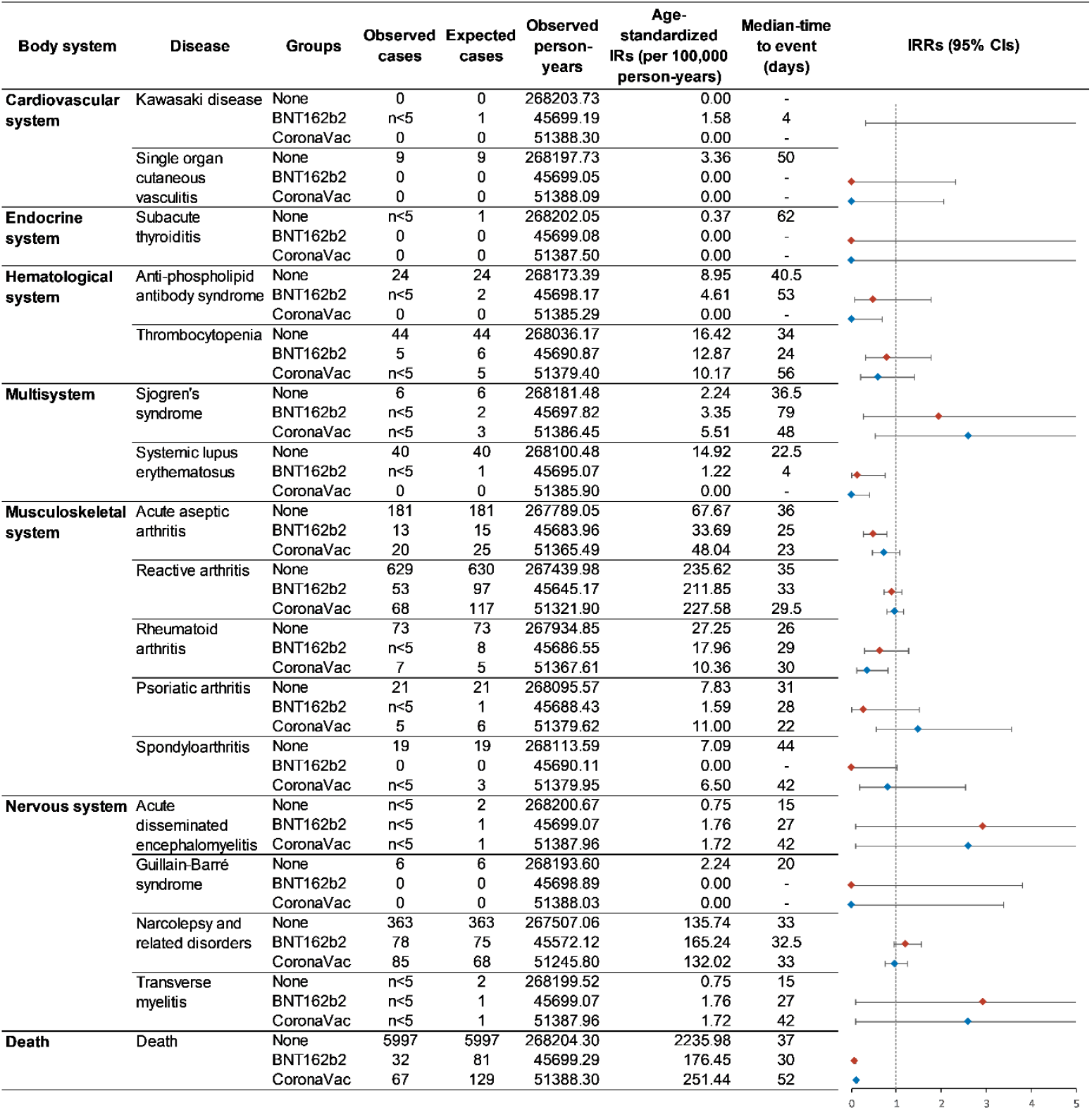
Standardized incidence rates up to the end date of data availability among second dose vaccine recipients and non-vaccinated individuals. Note: The line for CIs without a cap means the upper CI is greater than 5. Abbreviations: CIs, confidence intervals, IRs, incidence rates, IRRs, incidence rate ratios.

**Supplementary Figure 5.**
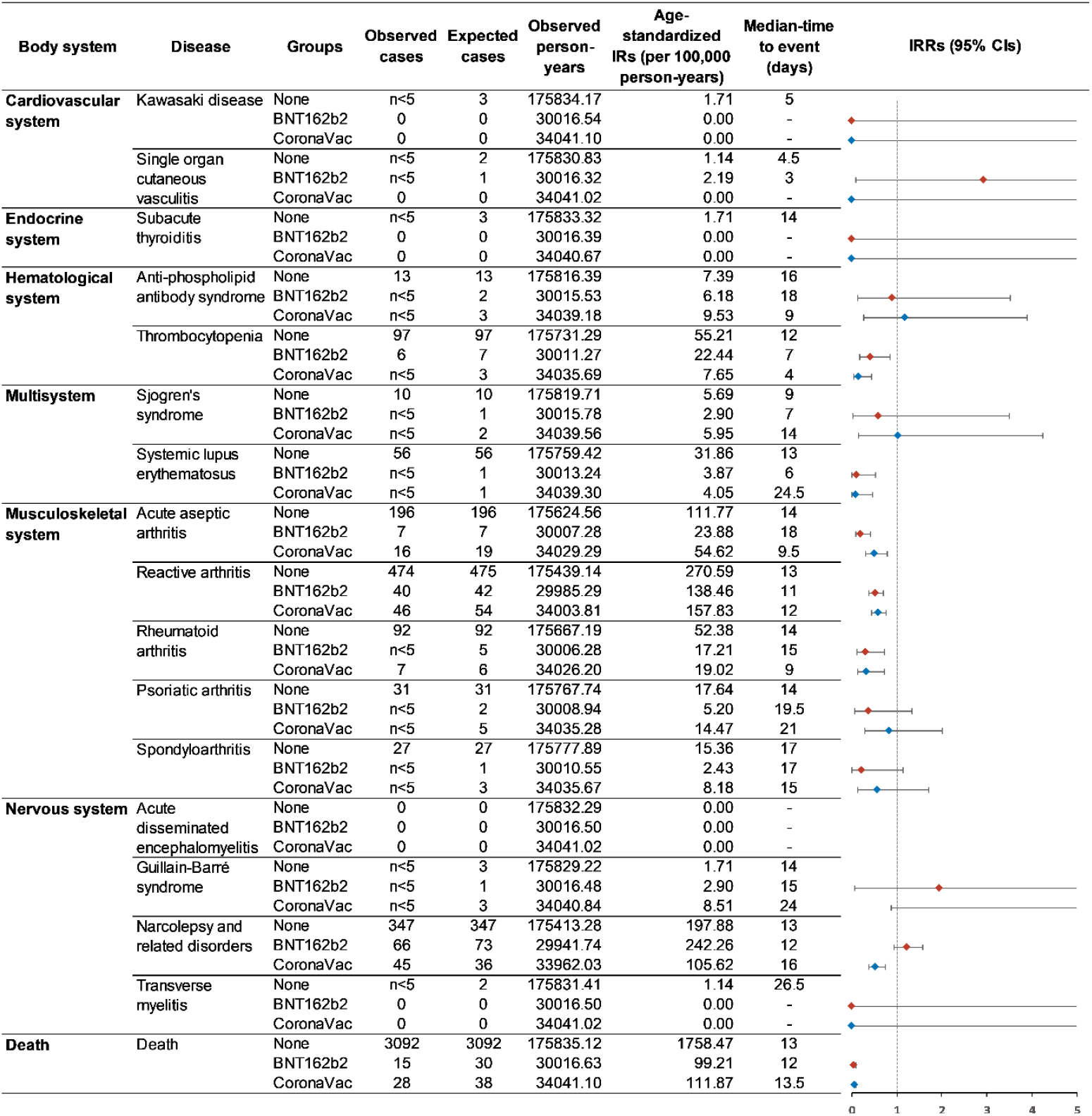
Standardized incidence rates of autoimmune conditions among first dose vaccine recipients and non-vaccinated individuals using both primary and secondary diagnosis for case identification. Note: The line for CIs without a cap means the upper CI is greater than 5. Abbreviations: CIs, confidence intervals, IRs, incidence rates, IRRs, incidence rate ratios.

**Supplementary Figure 6.**
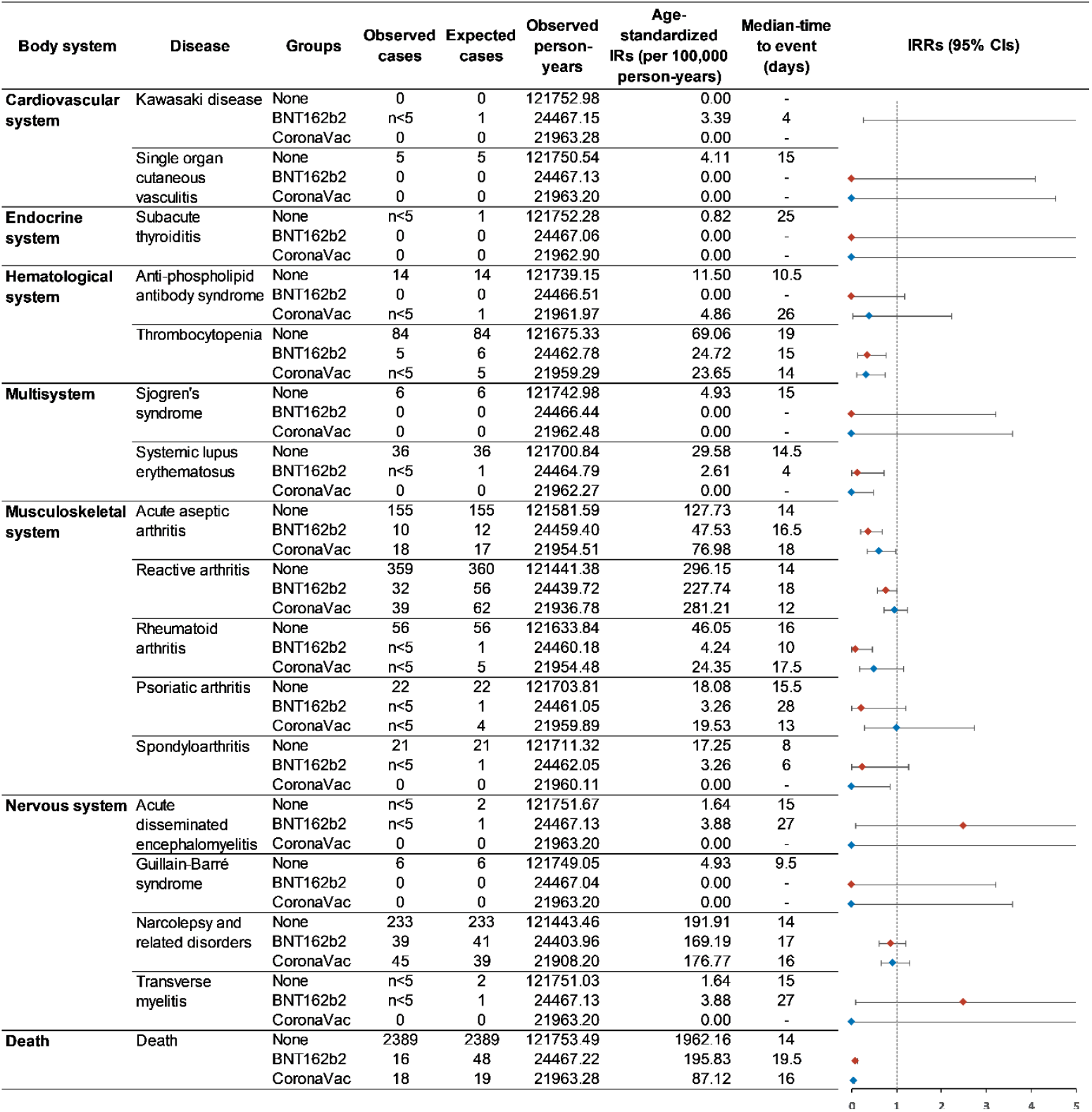
Standardized incidence rates of autoimmune conditions among second dose vaccine recipients and non-vaccinated individuals using both primary and secondary diagnosis for case identification. Note: The line for CIs without a cap means the upper CI is greater than 5. Abbreviations: CIs, confidence intervals, IRs, incidence rates, IRRs, incidence rate ratios.

**Supplementary Figure 7.**
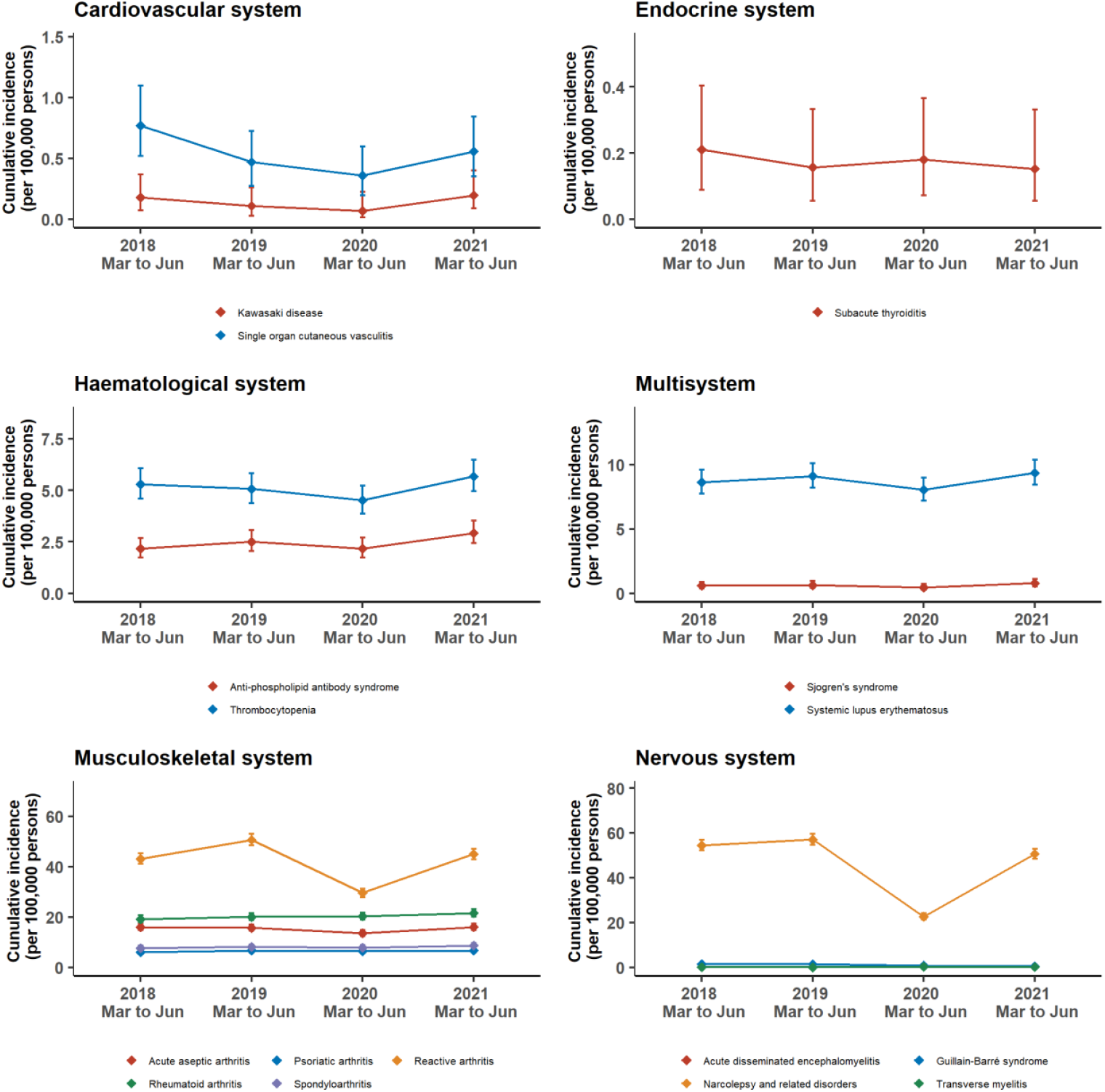
Trend of four-month cumulative incidence between 2018 and 2021.

**Supplementary Table 1.** ICD-9-CM diagnosis codes used for case identification.

| Body system | Autoimmune disease | ICD-9-CM | Description |
| --- | --- | --- | --- |
| Cardiovascular system | Kawasaki Disease<sup>1</sup> | 446.1 | Acute febrile mucocutaneous lymph node syndrome [MCLS] |
|  | Single Organ Cutaneous Vasculitis | 709.1 | Vascular disorders of skin |
|  |  | 446.2 | Hypersensitivity angiitis |
|  |  | 287.0 | Allergic purpura |
| Endocrine system | Subacute thyroiditis | 245.1 | Subacute thyroiditis |
| Hematological system | Anti-phospholipid antibody syndrome<sup>2-4</sup> | 795.79 | Other and unspecified nonspecific immunological findings |
|  |  | 286.5 | Hemorrhagic disorder due to intrinsic circulating anticoagulants |
|  |  | 289.8 | Other specified diseases of blood and blood-forming organs |
|  | Thrombocytopenia | 287.3 | Primary thrombocytopenia |
|  |  | 287.4 | Secondary thrombocytopenia |
|  |  | 287.5 | Thrombocytopenia, unspecified |
|  |  | 279.12 | Wiskott-Aldrich syndrome |
|  |  | 283.11 | Hemolytic-uremic syndrome |
|  |  | 284.1 | Pancytopenia |
|  |  | 446.6 | Thrombotic microangiopathy |
|  |  | 776.1 | Transient neonatal thrombocytopenia |
| Multisystem | Sjogren's syndrome | 710.2 | Sicca syndrome |
|  | Systemic lupus erythematosus | 710.0 | Systemic lupus erythematosus |
| Musculoskeletal system | Acute aseptic arthritis | 274.0 | Gouty arthropathy |
|  |  | 696.0 | Psoriatic arthropathy |
|  |  | 716.5 | Unspecified polyarthropathy or polyarthritis |
|  |  | 716.6 | Unspecified monoarthritis |
|  |  | 716.9 | Unspecified arthropathy |
|  |  | 712 | Crystal arthropathies |
|  |  | 711.5 | Arthropathy associated with other viral diseases |
|  | Reactive arthritis<sup>5,6</sup> | 099.3 | Reiter's disease |
|  |  | 711.1 | Arthropathy associated with Reiter's disease and nonspecific urethritis |
|  |  | 711.3 | Postdysenteric arthropathy |
|  |  | 716.4 | Transient arthropathy |
|  |  | 716.5 | Unspecified polyarthropathy or polyarthritis |
|  |  | 716.6 | Unspecified monoarthritis |
|  |  | 716.9 | Unspecified arthropathy |
|  |  | 719.4 | Pain in joint |
|  |  | 372.33 | Conjunctivitis in mucocutaneous disease |
|  | Rheumatoid arthritis | 714 | Rheumatoid arthritis and other inflammatory polyarthropathies |
|  | Psoriatic arthritis | 696 | Psoriasis and similar disorders |
|  | Spondyloarthritis | 720 | Ankylosing spondylitis and other inflammatory spondylopathies |
| Nervous system | Acute disseminated encephalomyelitis | 323.6 | Postinfectious encephalitis, myelitis, and encephalomyelitis |
|  |  | 323.8 | Other causes of encephalitis, myelitis, and encephalomyelitis |
|  | Guillain-Barré Syndrome | 357.0 | Acute infective polyneuritis |
|  |  | 357.8 | Other inflammatory and toxic neuropathy |
|  |  | 357.9 | Unspecified inflammatory and toxic neuropathy |
|  | Narcolepsy and related disorders<sup>7,8</sup> | 347 | Cataplexy and narcolepsy |
|  |  | 307.4 | Specific disorders of sleep of nonorganic origin |
|  |  | 780.5 | Sleep disturbances |
|  | Transverse myelitis | 341.2 | Acute (transverse) myelitis |
|  |  | 323.0 | Encephalitis, myelitis, and encephalomyelitis in viral diseases classified elsewhere |
|  |  | 323.4 | Other encephalitis, myelitis, and encephalomyelitis due to infection classified elsewhere |
|  |  | 323.5 | Encephalitis, myelitis, and encephalomyelitis following immunization procedures |
|  |  | 323.6 | Postinfectious encephalitis, myelitis, and encephalomyelitis |
|  |  | 323.8 | Other causes of encephalitis, myelitis, and encephalomyelitis |

Note: for those diseases without specific ICD-9-CM diagnosis code, series of ICD-9-CM diagnosis codes based on literature were used as approximated diagnosis. Abbreviation: ICD-9-CM, International Classification of Diseases, Ninth Revision, Clinical Modification.

**Supplementary Table 2.** Median-time to event (days, interquartile range) within 28 days after vaccination.

| Body system | Disease | First dose |  | Second dose |  |
| --- | --- | --- | --- | --- | --- |
|  |  | BNT162b2 | CoronaVac | BNT162b2 | CoronaVac |
| Cardiovascular system | Kawasaki disease | - | - | 4 (4-4) | - |
|  | Single organ cutaneous vasculitis | 3 (3-3) | - | - | - |
| Endocrine system | Subacute thyroiditis | - | - | - | - |
| Hematological system | Anti-phospholipid antibody syndrome | 18 (15.5-20.5) | 9 (3.75-14.75) | - | - |
|  | Thrombocytopenia | 4 (3-7.5) | 27 (27-27) | 21.5 (16.25-24.75) | - |
| Multisystem | Sjogren's syndrome | 7 (7-7) | 23 (23-23) | - | - |
|  | Systemic lupus erythematosus | 6 (6-6) | 24.5 (23.75-25.25) | 4 (4-4) | - |
| Musculoskeletal system | Acute aseptic arthritis | 11.5 (4.75-19.5) | 9.5 (5.5-17.5) | 16.5 (13-20.5) | 20.5 (11.5-23) |
|  | Reactive arthritis | 11 (7.75-14.5) | 10 (7-21.5) | 18 (9.25-21.5) | 11 (7-22) |
|  | Rheumatoid arthritis | 16.5 (13.75-19.25) | 9.5 (9-12.75) | 10 (9-11) | 19 (17.5-19.5) |
|  | Psoriatic arthritis | 19.5 (18.75-20.25) | 26.5 (25.75-27.25) | 28 (28-28) | 13 (10-17.5) |
|  | Spondyloarthritis | 17 (17-17) | 4 (4-4) | - | - |
| Nervous system | Acute disseminated encephalomyelitis | - | - | 27 (27-27) | - |
|  | Guillain-Barré syndrome | 15 (15-15) | - | - | - |
|  | Narcolepsy and related disorders | 13 (8.25-17.25) | 18.5 (11.5-22.75) | 17 (11.5-22.25) | 16 (11.5-21.5) |
|  | Transverse myelitis | - | - | 27 (27-27) | - |
| All-cause death | Death | 12 (8-17) | 13.5 (6.75-23.25) | 19.5 (13-23.25) | 16 (12-22) |

## References

1. Vellozzi C, Iqbal S, Broder K. Guillain-Barre syndrome, influenza, and influenza vaccination: the epidemiologic evidence. Clin Infect Dis 2014; 58(8): 1149–55.

2. Andrews N, Stowe J, Miller E, et al. A collaborative approach to investigating the risk of thrombocytopenic purpura after measles-mumps-rubella vaccination in England and Denmark. Vaccine 2012; 30(19): 3042–6.

3. Dubé E, Vivion M, MacDonald NE. Vaccine hesitancy, vaccine refusal and the anti-vaccine movement: influence, impact and implications. Expert Rev Vaccines 2015; 14(1): 99–117.

4. Creech CB, Walker SC, Samuels RJ. SARS-CoV-2 Vaccines. JAMA 2021; 325(13): 1318–20.

5. Baden LR, El Sahly HM, Essink B, et al. Efficacy and Safety of the mRNA-1273 SARS-CoV-2 Vaccine. N Engl J Med 2021; 384(5): 403–16.

6. Voysey M, Clemens SAC, Madhi SA, et al. Safety and efficacy of the ChAdOx1 nCoV-19 vaccine (AZD1222) against SARS-CoV-2: an interim analysis of four randomised controlled trials in Brazil, South Africa, and the UK. The Lancet 2021; 397(10269): 99–111.

7. Simpson CR, Shi T, Vasileiou E, et al. First-dose ChAdOx1 and BNT162b2 COVID-19 vaccines and thrombocytopenic, thromboembolic and hemorrhagic events in Scotland. Nature Medicine 2021; 27(7): 1290–7.

8. Wan EYF, Chui CSL, Lai FTT, et al. Bell’s palsy following vaccination with mRNA (BNT162b2) and inactivated (CoronaVac) SARS-CoV-2 vaccines: a case series and nested case-control study. The Lancet Infectious Diseases 2021; online first.

9. Hasan T, Khan M, Khan F, Hamza G. Case of Guillain-Barré syndrome following COVID-19 vaccine. BMJ Case Reports 2021; 14(6): e243629.

10. Maramattom BV, Krishnan P, Paul R, et al. Guillain-Barré Syndrome following ChAdOx1-S/nCoV-19 Vaccine. Annals of Neurology; n/a(n/a).

11. Ogbebor O, Seth H, Min Z, Bhanot N. Guillain-Barré syndrome following the first dose of SARS-CoV-2 vaccine: A temporal occurrence, not a causal association. IDCases 2021; 24: e01143.

12. Finsterer J. Exacerbating Guillain–Barré Syndrome Eight Days after Vector-Based COVID-19 Vaccination. Case Reports in Infectious Diseases 2021; 2021: 3619131.

13. McMahon DE, Amerson E, Rosenbach M, et al. Cutaneous reactions reported after Moderna and Pfizer COVID-19 vaccination: A registry-based study of 414 cases. J Am Acad Dermatol 2021; 85(1): 46–55.

14. An QJ, Qin DA, Pei JX. Reactive arthritis after COVID-19 vaccination. Hum Vaccin Immunother 2021: 1–3.

15. Watad A, De Marco G, Mahajna H, et al. Immune-Mediated Disease Flares or New-Onset Disease in 27 Subjects Following mRNA/DNA SARS-CoV-2 Vaccination. Vaccines (Basel) 2021; 9(5): 435.

16. Chan EW, Lau WCY, Leung WK, et al. Prevention of Dabigatran-Related Gastrointestinal Bleeding With Gastroprotective Agents: A Population-Based Study. Gastroenterology 2015; 149(3): 586-95.e3.

17. Lau WCY, Chan EW, Cheung C-L, et al. Association Between Dabigatran vs Warfarin and Risk of Osteoporotic Fractures Among Patients With Nonvalvular Atrial Fibrillation. JAMA 2017; 317(11): 1151–8.

18. Man KKC, Coghill D, Chan EW, et al. Association of Risk of Suicide Attempts With Methylphenidate Treatment. JAMA Psychiatry 2017; 74(10): 1048–55.

19. Wong AYS, Root A, Douglas IJ, et al. Cardiovascular outcomes associated with use of clarithromycin: population based study. BMJ 2016; 352: h6926.

20. Cohen GR, Yang SY. Mid-P confidence intervals for the Poisson expectation. Stat Med 1994; 13(21): 2189–203.

21. Fay MP. Two-sided Exact Tests and Matching Confidence Intervals for Discrete Data. The R Journal 2010; 2(1): 53–8.

22. Kharkar V, Vishwanath T, Mahajan S, Joshi R, Gole P. Asymmetric cutaneous vasculitis following COVID-19 vaccination with unusual eosinophil preponderance. Clin Exp Dermatol 2021.

23. The World Health Organization. Covid-19 vaccines: safety surveillance manual. 2020. https://www.who.int/publications/i/item/10665338400 (accessed July 25 2021).

24. Pottegård A, Lund LC, Karlstad Ø, et al. Arterial events, venous thromboembolism, thrombocytopenia, and bleeding after vaccination with Oxford-AstraZeneca ChAdOx1-S in Denmark and Norway: population based cohort study. BMJ 2021; 373: n1114.

25. Menni C, Klaser K, May A, et al. Vaccine side-effects and SARS-CoV-2 infection after vaccination in users of the COVID Symptom Study app in the UK: a prospective observational study. The Lancet Infectious Diseases 2021; 21(7): 939–49.

26. The Government of the Hong Kong Special Administrative Region. Vaccination priority groups to be expanded to cover people aged 30 or above. 2021. https://www.info.gov.hk/gia/general/202103/15/P2021031500626.htm.

27. Gadroen K, Straus S, Pacurariu A, Weibel D, Kurz X, Sturkenboom M. Patterns of spontaneous reports on narcolepsy following administration of pandemic influenza vaccine; a case series of individual case safety reports in Eudravigilance. Vaccine 2016; 34(41): 4892–7.

28. Granath F, Gedeborg R, Smedje H, Feltelius N. Change in risk for narcolepsy over time and impact of definition of onset date following vaccination with AS03 adjuvanted pandemic A/H1N1 influenza vaccine (Pandemrix) during the 2009 H1N1 influenza pandemic. Pharmacoepidemiol Drug Saf 2019; 28(8): 1045–53.

29. Montplaisir J, Petit D, Quinn MJ, et al. Risk of narcolepsy associated with inactivated adjuvanted (AS03) A/H1N1 (2009) pandemic influenza vaccine in Quebec. PLoS One 2014; 9(9): e108489.

30. O’Flanagan D, Barret AS, Foley M, et al. Investigation of an association between onset of narcolepsy and vaccination with pandemic influenza vaccine, Ireland April 2009-December 2010. Euro Surveill 2014; 19(17): 15–25.

31. Stowe J, Andrews N, Kosky C, et al. Risk of Narcolepsy after AS03 Adjuvanted Pandemic A/H1N1 2009 Influenza Vaccine in Adults: A Case-Coverage Study in England. Sleep 2016; 39(5): 1051–7.

32. Winstone AM, Stellitano L, Verity C, et al. Clinical features of narcolepsy in children vaccinated with AS03 adjuvanted pandemic A/H1N1 2009 influenza vaccine in England. Developmental Medicine & Child Neurology 2014; 56(11): 1117–23.

33. Narcolepsy fiasco spurs Covid vaccine fears in Sweden. 2020. https://medicalxpress.com/news/2020-11-narcolepsy-fiasco-spurs-covid-vaccine.html (accessed July 22, 2021).

34. Silberner J. Overcoming vaccine hesitancy: five minutes with Heidi Larson. BMJ 2019; 364: l1259.

35. Wei Y-T, Lee P-Y, Lin C-Y, et al. Non-alcoholic fatty liver disease among patients with sleep disorders: a Nationwide study of Taiwan. BMC Gastroenterology 2020; 20(1): 32.

36. VAC4EU COVID-19 vaccine monitoring. 2021. https://vac4eu.org/covid-19-vaccine-monitoring/ (accessed July 28, 2021).

37. Hajduczok AG, DiJoseph KM, Bent B, et al. Physiologic Response to the Pfizer-BioNTech COVID-19 Vaccine Measured Using Wearable Devices: A Prospective Observational Study. JMIR Form Res 2021.

## References

1. Baker MA, Baer B, Kulldorff M, et al. Kawasaki disease and 13-valent pneumococcal conjugate vaccination among young children: A self-controlled risk interval and cohort study with null results. PLOS Medicine 2019; 16(7): e1002844.

2. Algahtani FH, AlQahtany FS, ElGohary G, et al. The clinical and laboratory manifestations profile of antiphospholipid syndrome among Saudi Arabia population: Examining the applicability of Sapporo criteria. Saudi Journal of Biological Sciences 2020; 27(9): 2425–30.

3. Somers EC, Marder W, Cagnoli P, et al. Population-based incidence and prevalence of systemic lupus erythematosus: the Michigan Lupus Epidemiology and Surveillance program. Arthritis Rheumatol 2014; 66(2): 369–78.

4. Chen H-H, Lin C-H, Chao W-C. Risk of Systemic Lupus Erythematosus in Patients With Anti-phospholipid Syndrome: A Population-Based Study. Frontiers in Medicine 2021; 8(559).

5. Curry JA, Riddle MS, Gormley RP, Tribble DR, Porter CK. The epidemiology of infectious gastroenteritis related reactive arthritis in U.S. military personnel: a case-control study. BMC Infectious Diseases 2010; 10(1): 266.

6. Australian Governement. Reactive Arthritis A006. 29 October 2018. https://clik.dva.gov.au/ccps-medical-research-library/sops-grouped-icd-body-system/q-z/reactive-arthritis-a006-m02.

7. Background rates of Adverse Events of Special Interest for monitoring COVID-19 vaccines. Full protocol. http://www.encepp.eu/encepp/openAttachment/fullProtocol/37296.

8. Wei YT, Lee PY, Lin CY, et al. Non-alcoholic fatty liver disease among patients with sleep disorders: a Nationwide study of Taiwan. BMC gastroenterology. 2020; 20(1): 1–8.

